# Decision Curve Analysis for Evaluating Machine Learning Models for Next-Day Transfer Out of ICU

**DOI:** 10.64898/2026.04.19.26351213

**Authors:** Margaret Pozo, Abigail Pape, Brian Locke, Warren Woodrich Pettine

**Author notes:** Co-senior author. Corresponding author. Co-senior author.

## Abstract

Timely identification of intensive care unit (ICU) patients likely to exit the unit can support anticipatory workflows such as chart review, eligibility screening, and patient outreach prior to transfer. Most ICU discharge prediction studies report discrimination and calibration, but these metrics do not quantify the decision consequences of acting on predictions. Using adult ICU admissions from MIMIC-IV, we represented each ICU stay as a sequence of daily clinical summaries and trained logistic regression, random forest, and XGBoost models to predict next day ICU transfer. Models achieved ROC AUC of 0.80–0.84 with differing calibration. We evaluated decision utility using decision curve analysis (DCA), where positive predictions trigger proactive review. Across thresholds, model guided strategies outperformed review-all, review-none, and a simple clinical rule. To translate net benefit into implementable operations, we modeled a clinical trial recruitment workflow with an 8 hour daily time constraint, incorporating chart review and consent effort. At a feasible operating threshold (0.23), the model flagged ∼23 charts/day and yielded ∼1.23 enrollments/day under conservative eligibility and consent assumptions. These results demonstrate that DCA provides a transparent framework for determining when ICU transfer predictions are worth using and how thresholds should be selected to align with real world workflow constraints.

**Data and Code Availability:** This research has been conducted using data from MIMIC-IV. Researchers can request access via PhysioNet. Implementation code is available upon request.

## 1. Introduction

In the intensive care unit (ICU), many downstream activities, such as chart review, eligibility screening for clinical trials, and preparatory co-ordination for post-ICU care, must be completed shortly before a patient leaves the unit. Research staff often lack reliable signals identifying which patients are likely to be discharged the following day, leading to ad hoc judgment or universal screening that strains limited personnel resources Rubins and Moskowitz (1988); Franklin and Jackson (1983).

Clinical trial recruitment is a major driver of cost and delay in drug development, particularly in acute care settings where eligibility windows are short and screening must occur rapidly Martin et al. (2017); Khanna (2012); Gul and Ali (2010); Chaudhari et al. (2020). Inefficient or poorly targeted review in the ICU can therefore meaningfully affect trial feasibility, staffing burden, and cost, motivating the use of discharge prediction models as tools for prioritizing recruitment effort rather than forecasting alone.

A growing body of research has proposed machine learning (ML) models to predict discharge timing, length of stay, or readmission risk using electronic health record (EHR) data Verburg et al. (2017); Rojas et al. (2018); Badawi and Breslow (2012). Often these reportings include strong discrimination and calibration using metrics such as ROC AUC or calibration slope. However, predictive accuracy alone does not determine practical usefulness. High discrimination does not indicate whether acting on predictions improves decision making when actions carry asymmetric costs, such as unnecessary chart review versus missed opportunities to identify imminently discharged patients Vickers et al. (2019); Vickers and Holland (2021); Kappen et al. (2018). Without explicit linkage between predictions and decisions, it remains unclear how probability thresholds should be interpreted or how models compare to simple non-model strategies, leaving clinicians and research staff without actionable guidance Antoniou and Mamdani (2021).

Decision curve analysis (DCA) provides a principled framework for addressing this gap by evaluating models based on net benefit across decision thresholds. Rather than asking which model discriminates best, DCA asks whether using a model improves outcomes relative to baseline strategies such as acting on all patients or none. In this study, we frame next day ICU transfer prediction as a decision support problem for prioritizing proactive review in clinical trial recruitment. Using adult ICU admissions from MIMIC-IV, we construct daily patient representations from end-of-day data and evaluate common supervised learning models. By extending DCA to incorporate realistic staffing and time constraints, we show when discharge timing predictions meaningfully improve prioritization and when apparent gains reflect near universal flagging rather than operational benefit.

## 2. Methods

We used a retrospective prognostic modeling framework based on patient day observations within ICU admissions. The prediction task was to classify whether an ICU patient would transfer from the ICU to the hospital floor within the subsequent 24 hours, using only information available up to midnight of the current calendar day (Figure 1). For a given calendar day D, features were constructed from observations recorded up to 23:59 on day D, and the binary outcome indicated whether ICU transfer occurred at any time during day D+1. This temporal alignment ensured that all predictors preceded the outcome and prevented leakage of future information into model training or evaluation.

**Figure 1:**
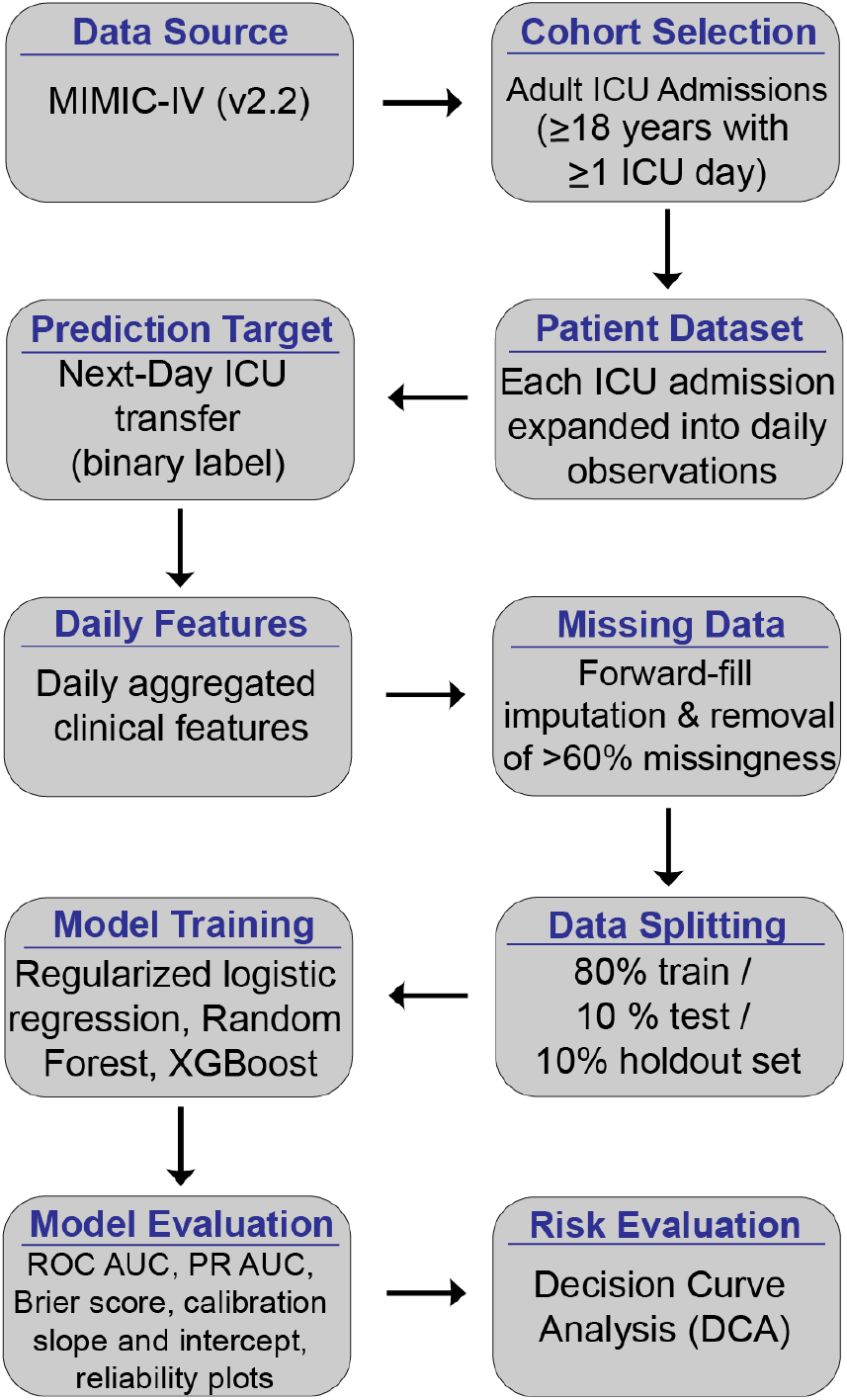
Modeling pipeline for next-day ICU transfer prediction. Overview of the pipeline used to construct, train, and evaluate next day ICU transfer using discrimination, calibration, and decision curve analysis.

Data were obtained from the MIMIC-IV (v2.2) database, which contains de identified EHR data for adult ICU admissions at a large academic medical center from 2008 to 2019. The database includes demographics, time stamped vital signs, laboratory measurements, interventions, and ICU transfer events. The cohort included adult patients (≥18 years) with at least one ICU admission and at least one recorded physiologic or laboratory measurement. ICU stays shorter than 24 hours, admissions with missing or ambiguous ICU exit times, and implausible ICU durations were excluded. Each ICU admission was expanded into a sequence of patient day observations.

All features were aggregated at the patient day level using data available up to the end of each calendar day. Features were organized into five domains: vital signs, laboratory measurements, early warning scores, interventions, and ICU trajectory. Vital sign features included daily minimum, mean, and standard deviation for heart rate, respiratory rate, systolic blood pressure, and temperature, along with short term temporal trends computed over 6- or 12-hour windows to capture physiologic stabilization or deterioration. Laboratory features reflected renal, metabolic, and hematologic status, including creatinine, blood urea nitrogen, lactate, bicarbonate, pH, electrolytes, white blood cell count, and hemoglobin; for each variable, the daily mean, most recent value, and within day variability were extracted. Modified Early Warning Scores (MEWS) were computed using values available up to that calendar day. Intervention features captured mechanical ventilation status and settings (FiO_2_, PEEP), vasopressor use (binary indicators and norepinephrine equivalent dose), and renal replacement therapy. ICU trajectory features included current ICU day and cumulative ICU length of stay.

Clinical data completeness varied across features and reflected both measurement frequency and clinical practice patterns. To preserve temporal continuity without introducing future information, missing values within an ICU admission were forward filled using the most recent prior observation. For variables that were never observed during a given admission, median imputation was applied using statistics computed from the training data only. Features with more than 60% missingness across patient days were excluded to reduce reliance on imputation and limit potential bias. After filtering and preprocessing, the final dataset contained approximately 78,000 patient days and 215 features.

To prevent patient overlap, all data splits were performed at the ICU admission level. Admissions were partitioned into training (81%), internal test (9%), and final holdout (10%) sets. Hyperparameters were tuned using 10-fold stratified cross validation within the training set. All reported performance metrics and decision analyses were computed on the internal test set unless otherwise stated; the final holdout set was reserved for confirmation analyses.

Three supervised learning models were evaluated: L2 regularized logistic regression, random forest, and extreme gradient boosting (XG-Boost). No class reweighting or resampling was applied in order to preserve the natural outcome prevalence. All preprocessing steps, including imputation, scaling, and one hot encoding, were embedded within each model pipeline and fitted only on training folds.

Predictive performance was assessed using ROC AUC and precision–recall AUC. Calibration was evaluated using the Brier score, calibration intercept and slope from logistic recalibration, and 20 bin reliability plots. For tree based models, global feature contributions were assessed using SHAP values computed with the TreeExplainer framework.

Decision level utility was evaluated using DCA. In this framework, a positive prediction corresponds to initiating proactive chart review and eligibility screening on the following day, rather than recommending discharge or taking no action. Net benefit was computed across the full range of threshold probabilities *t* ∈ (0, 1) and compared against baseline strategies of reviewing all patient days, reviewing none, and a simple clinical heuristic Kappen et al. (2018).

Net benefit was defined as:

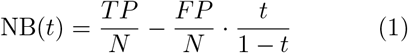

where TP and FP denote the number of true and false positive review decisions at threshold t, and N is the total number of patient days. In this formula, the threshold probability t encodes the relative tolerance for unnecessary review versus missed next day ICU transfers.

The clinical heuristic flagged a patient day for review if heart rate *<*100 bpm, respiratory rate *<*28 breaths/min, and no vasopressor therapy was administered in the prior 24 hours, reflecting a simple physiologic stability screen used as a non-model prioritization baseline.

To translate decision level net benefit into operational consequences, we instantiated a clinical recruitment workflow reflecting a resource constrained research setting. We assumed a fixed ICU census of 30 patient days per day and an 8 hour daily time budget (480 minutes) for a research coordinator responsible for chart review and recruitment.

For a given threshold t, predicted probabilities were binarized to generate review decisions. Let TP, FP, FN, and TN denote confusion matrix counts over the evaluation set. These quantities were converted to rates per 100 ICU patient days and scaled to daily counts using the assumed census. Review workload was computed as:

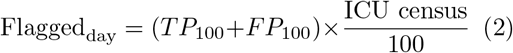

Chart review time was computed assuming a fixed review time of 15 minutes per flagged patient day. Recruitment time was computed only for eligible true positives, assuming a fixed eligibility rate among true transfers and a fixed time per approach and consent attempt (60 minutes). Total daily time was given by:

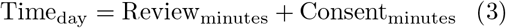

Thresholds were considered feasible if both the total daily time did not exceed 480 minutes and the number of flagged charts did not exceed the maximum daily review capacity.

To assess economic implications, we defined a simple net value function:

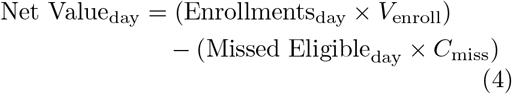

where enrollment yield depended on the assumed eligibility rate among true positives and consent success rate. All operational and economic parameters were chosen conservatively and treated as illustrative; sensitivity analyses across these parameters are reported. This framework allowed identification of threshold specific operating points that maximize expected value while satisfying staffing and time constraints.

## 3. Results

The final analytic cohort comprised approximately 78,000 ICU patient days derived from adult ICU admissions in MIMIC-IV. Among patient days labeled as next-day ICU exit (positive class), 76.89% exited to the hospital floor and 23.01% resulted in death or direct discharge home (Figure 2A). This high prevalence of next day ICU exit establishes a setting in which baseline strategies such as reviewing all patients are expected to perform strongly, emphasizing the need for decision focused evaluation rather than discrimination alone.

**Figure 2:**
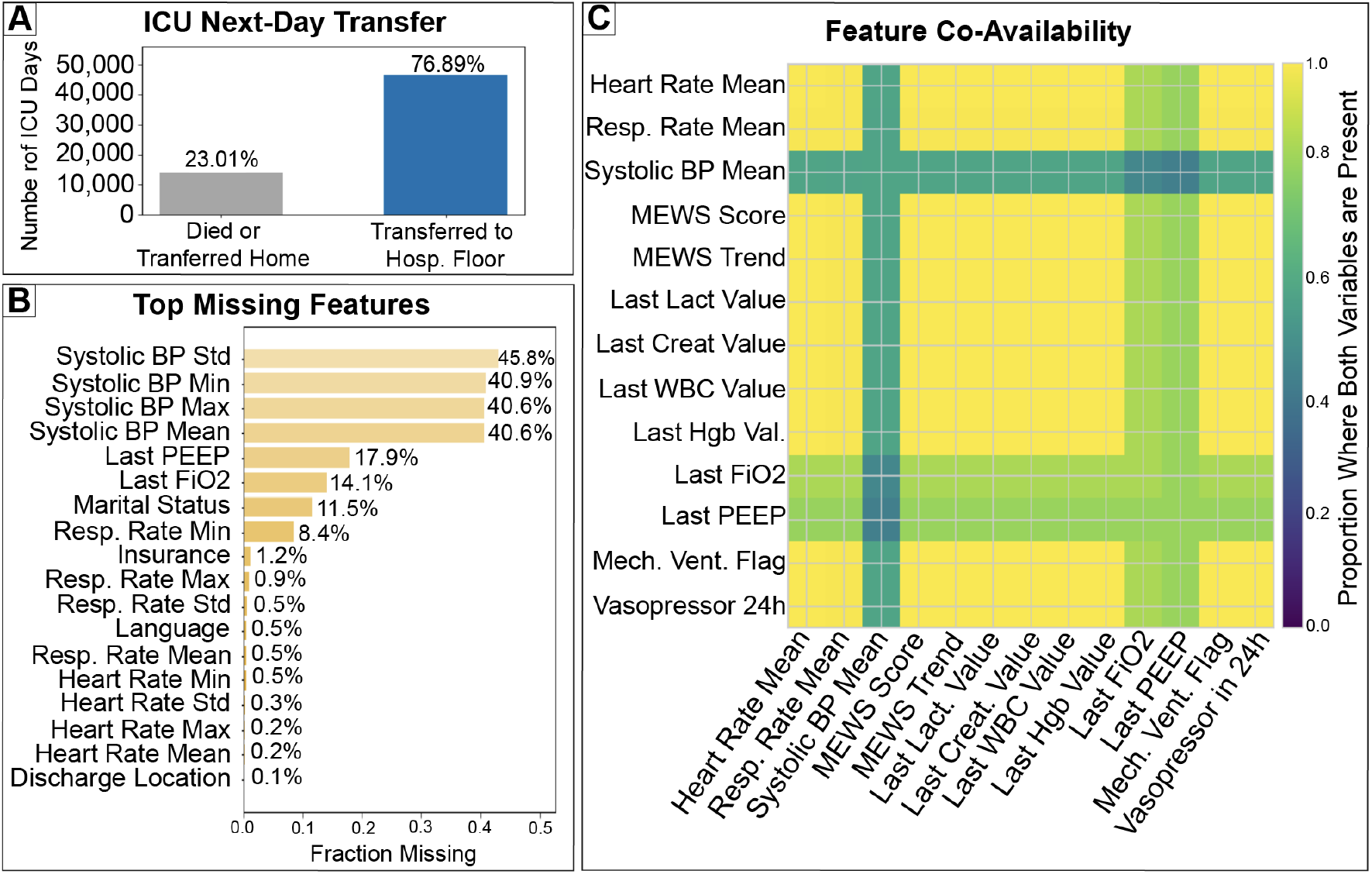
Next-day ICU transfers are common and characterized by heterogeneous observed clinical data. (A) Distribution of ICU patient days by next-day outcome, showing that most transfers occur to the hospital floor. (B) Fraction of missing values across selected features, illustrating variability in clinical measurement frequency. (C) Feature co-availability (overlap) across clinical domains, highlighting that multivariate information is available on most patient days despite heterogeneous measurement frequency.

The dataset contained approximately 78,134 patient days derived from 94,444 ICU stays. The median ICU length of stay was 2.1 days, and the patient day level outcome prevalence of next day ICU exit was 75.67% on the internal test set. Clinical data availability varied substantially across features. Measures derived from systolic blood pressure exhibited the highest missingness, with 45.8% missingness for systolic BP standard deviation and approximately 40–41% missingness for systolic BP minimum, maximum, and mean (Figure 2B). Intervention-related variables such as PEEP (17.9%) and FiO_2_ (14.1%) were moderately missing, while most vital signs and demographic variables had minimal missingness (*<* 1%). Despite heterogeneous measurement frequency, feature co-availability was high across most physiologic, laboratory, and intervention variables, with notable exceptions for systolic blood pressure summaries (≈ 0.4) and moderate co-availability for ventilatory parameters (≈ 0.8 for FiO_2_ and PEEP; Figure 2C). Together, these patterns indicate that although individual features are inconsistently measured, sufficient multivariate information is available on most patient days to support predictive modeling.

Results are reported on the internal test set unless otherwise noted. All evaluated models demonstrated moderate to strong discrimination for next day ICU transfer. ROC AUC ranged from 0.80 for logistic regression to 0.84 for random forest, with XGBoost achieving intermediate performance (Figure 3A). Precision–recall performance was similarly high across models, with PR AUC values exceeding 0.91 (Figure 3B), reflecting the high baseline prevalence of next day ICU exit.

**Figure 3:**
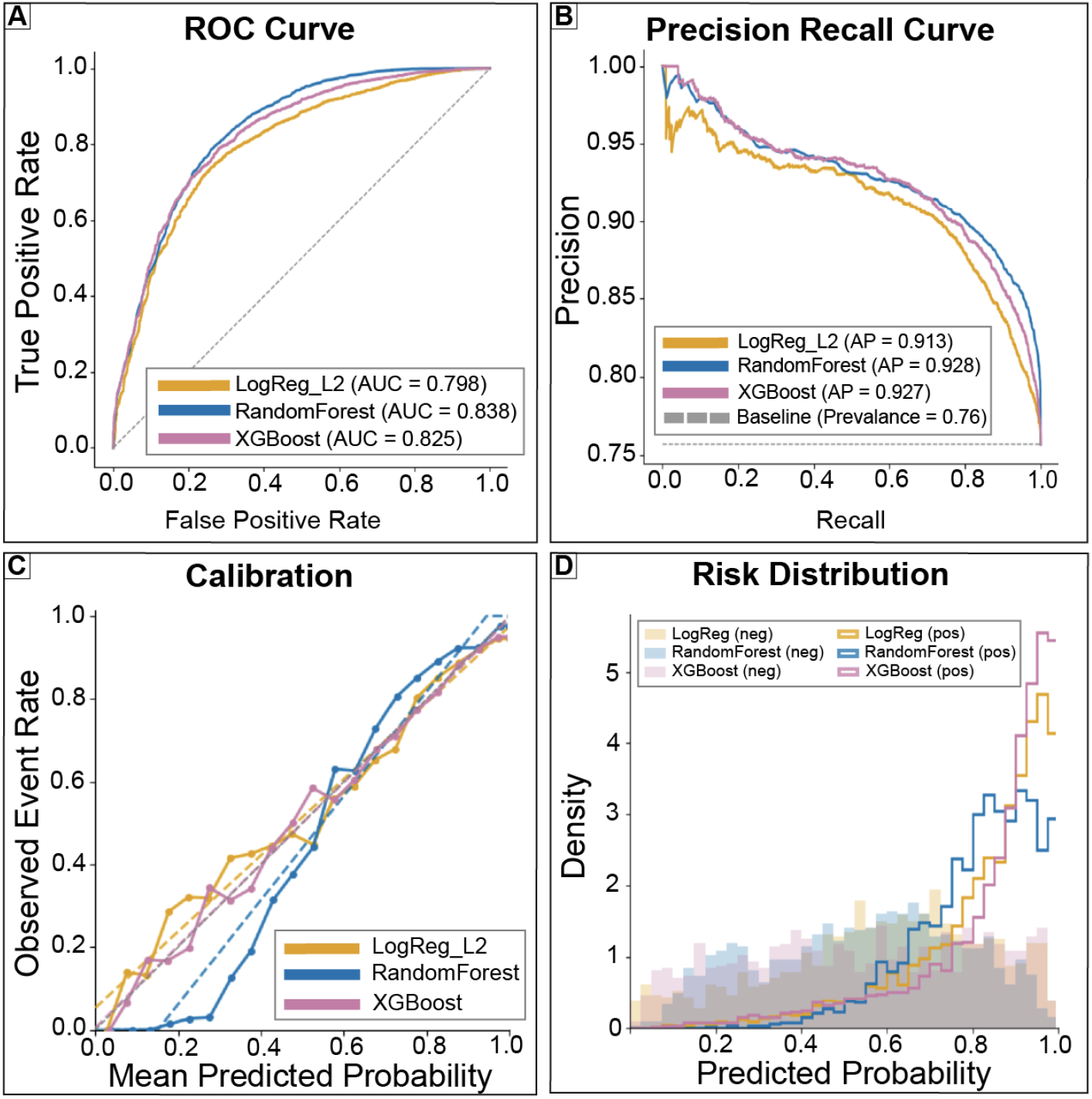
Comparable discrimination across models with differing calibration characteristics. (A) Receiver operating characteristic curves. (B) Precision–recall curves with baseline prevalence shown for reference. (C) Calibration curves comparing predicted probabilities to observed event rates. (D) Distributions of predicted risk scores stratified by outcome. Together, these panels summarize predictive behavior relevant to threshold based decision analysis.

Calibration characteristics differed meaningfully across models. Logistic regression and XG-Boost exhibited calibration slopes closer to unity (0.90 and 0.92, respectively), whereas random forest showed a steeper slope (1.40) and a negative calibration intercept, indicating systematic overestimation of risk at higher predicted probabilities (Figure 3C). Risk distribution plots revealed substantial overlap between discharged and non-discharged patient days, particularly at intermediate predicted probabilities (Figure 3D). Taken together, these findings show that while models rank patients similarly, calibration differences and overlapping risk distributions limit the interpretability of predicted probabilities without explicit threshold based decision analysis.

Feature importance patterns varied across modeling approaches (Figure 4). Logistic regression emphasized laboratory deltas and variability measures, such as changes in chloride and hematocrit, alongside admission characteristics and language indicators. In contrast, tree based models prioritized contemporaneous physiologic state and ICU trajectory, including ventilatory parameters (PEEP, FiO_2_), lactate, creatinine, current ICU day, and mechanical ventilation status. These differences suggest that models encode distinct representations of patient readiness for ICU transfer rather than converging on a single dominant predictor set, reinforcing the need to evaluate models based on decision level consequences rather than feature rankings alone.

**Figure 4:**
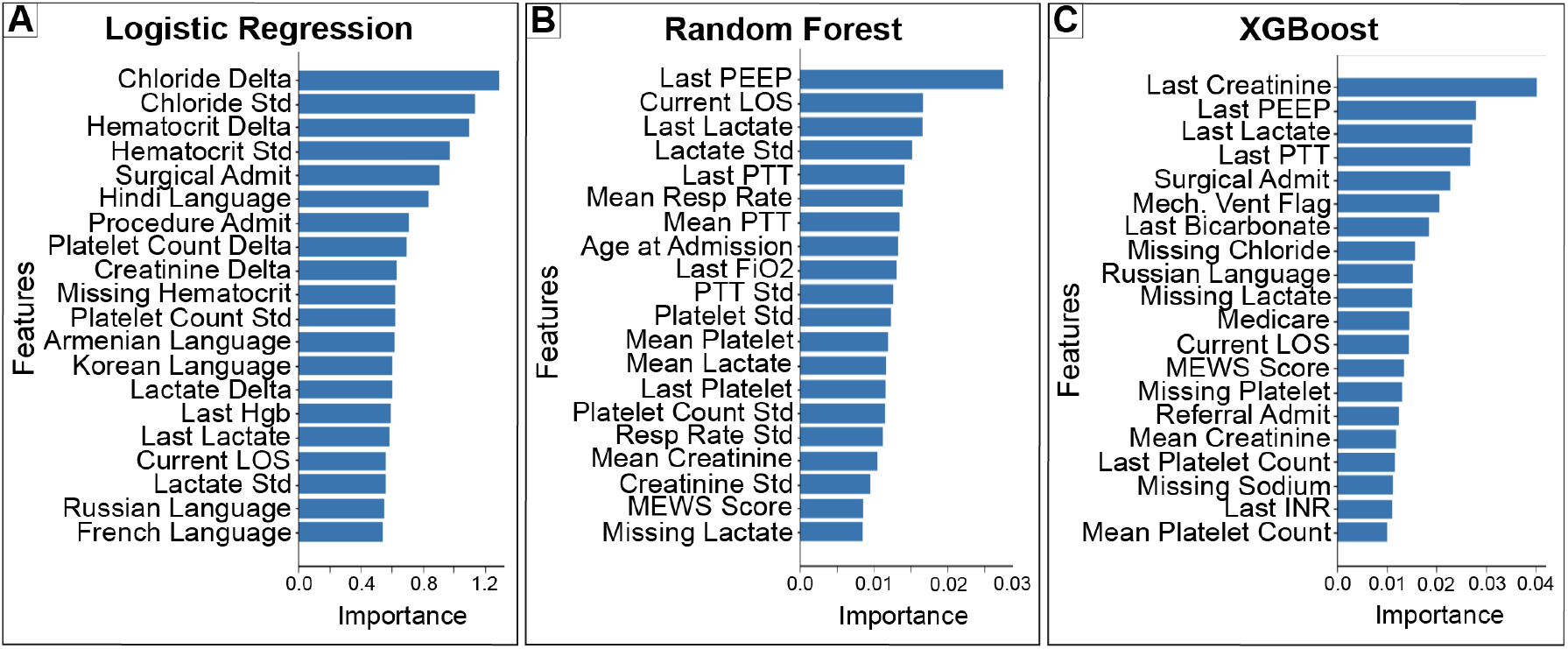
Model specific feature importance reflects representations of next day ICU discharge. (A) Regularized logistic regression coefficients (absolute magnitude). (B) Random forest feature importance based on impurity reduction. (C) XGBoost feature importance derived from split gain. Feature rankings differ across models, reflecting distinct representations of clinical information rather than a single dominant predictor set.

DCA demonstrated that across a broad range of thresholds, model guided review strategies achieved higher or comparable net benefit relative to review-all, review-none, and a simple clinical heuristic (Figure 5A). However, in this high prevalence setting, gains at low thresholds were often driven by near universal flagging, underscoring that favorable net benefit alone does not imply operational advantage. As the decision threshold increased, the fraction of patient days flagged for review declined monotonically (Figure 5B), accompanied by corresponding reductions in both true positives and false positives per 1,000 ICU days (Figure 5C–D). Table A summarizes representative thresholds (0.1–0.9), their implied false positive penalty weights, and their interpretation in terms of tolerance for unnecessary review versus missed next day ICU exits. Together, these results clarify how probability thresholds encode implicit cost ratios and why threshold selection must be informed by downstream constraints rather than predictive performance alone.

**Table 1:** Predictive performance and calibration of next day ICU transfer. Discrimination, calibration, and threshold specific performance metrics for supervised learning models predicting next day ICU discharge. Reported values include ROC AUC, precision–recall AUC, Brier score, F1 score at a fixed 0.5 threshold, and calibration intercept and slope estimated using logistic recalibration.

| Model | ROC AUC | PR AUC | Brier Score | F1 at 0.5 | Calibration |  |
| --- | --- | --- | --- | --- | --- | --- |
|  |  |  |  |  | Intercept | Slope |
| LogReg_L2 | 0.7982 | 0.9127 | 0.141 982 | 0.8723 | 0.0667 | 0.8952 |
| Random Forest | 0.8376 | 0.9281 | 0.125 025 | 0.9001 | −0.2348 | 1.3950 |
| XGBoost | 0.8252 | 0.9266 | 0.139 855 | 0.8855 | 0.0275 | 0.9158 |

**Figure 5:**
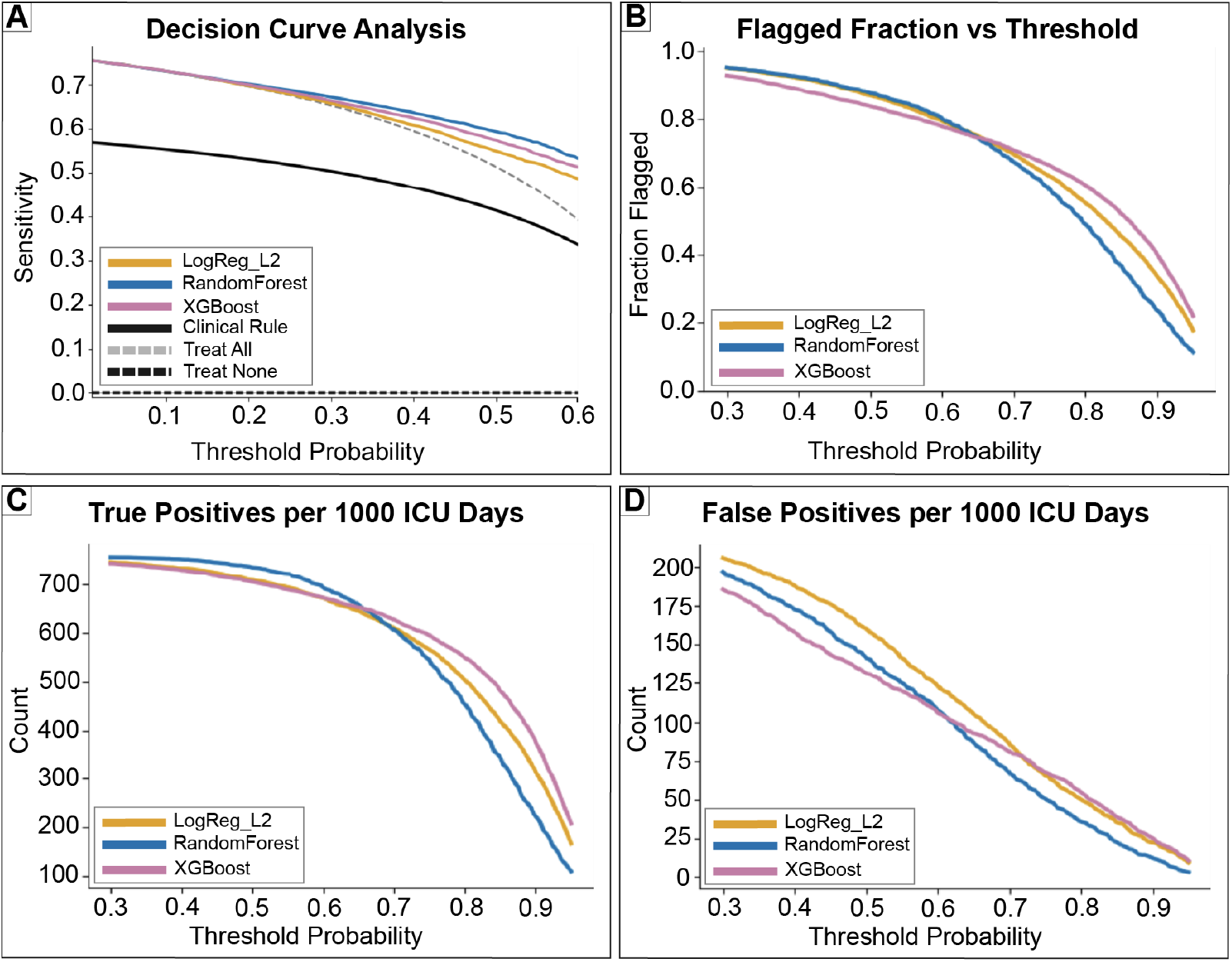
Decision curve analysis shows consistent net benefit across clinically plausible thresholds. (A) Decision curve analysis comparing model guided strategies to treat-all, treat-none, and a simple clinical rule. (B) Fraction of patient days flagged for review as a function of the decision threshold. (C) True positives per 1,000 ICU patient days. (D) False positives per 1,000 ICU patient days. Together, these panels quantify how threshold choice trades review burden against missed next day discharges.

To translate decision level net benefit into realistic trial operations, we evaluated model predictiocns within a constrained ICU clinical trial recruitment scenario incorporating coordinator time limits and a multistage enrollment funnel. Chart review time was set to 7 minutes per flagged patient day, approach and consent time to 60 minutes per eligible patient, and total coordinator availability to 8 hours per day Siems et al. (2020); Tew et al. (2023). Total daily coordinator workload varied nonlinearly with the decision threshold, reflecting the opposing effects of reduced chart review and increased missed eligible patients as thresholds increased (Figure 6A). As expected, increasing the threshold reduced chart review burden. However, once sufficient true positives were identified, total workload was increasingly dominated by approach and consent effort, leading to diminishing returns in total time savings at higher thresholds while increasing missed eligible patients.

**Figure 6:**
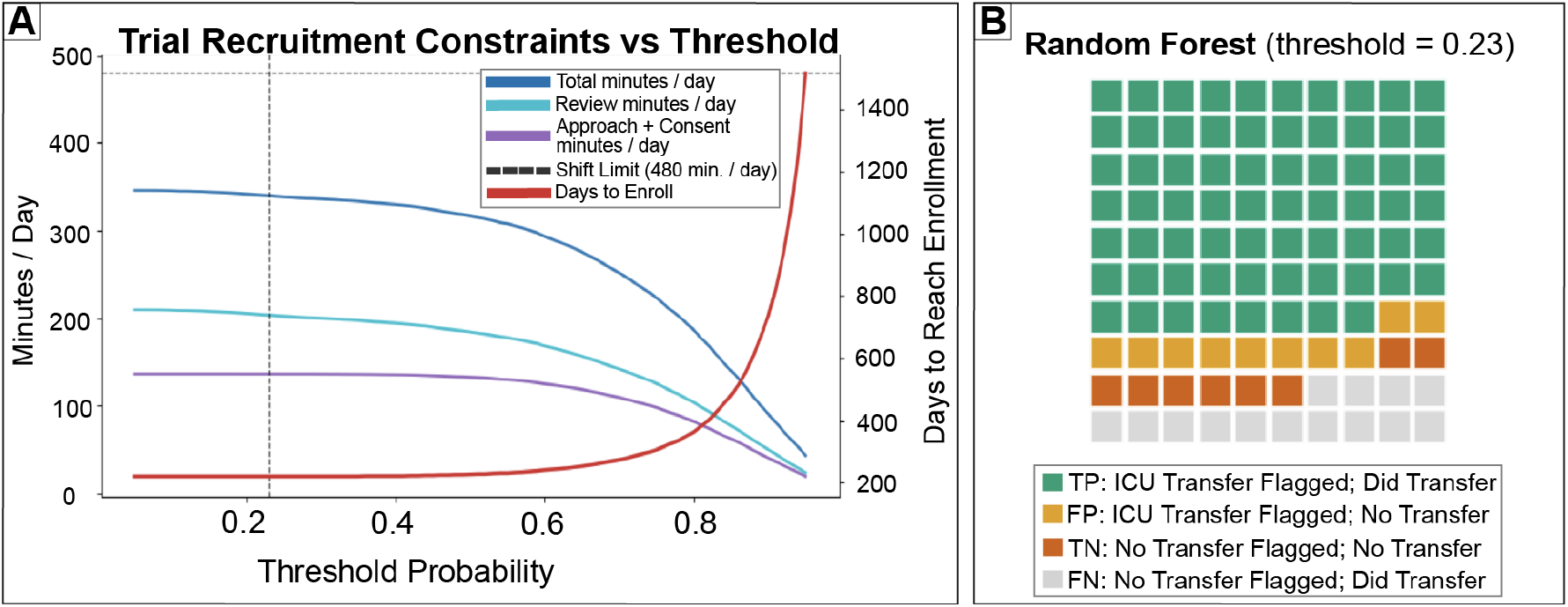
Operational feasibility analysis identifies an implementable decision threshold. (A) Daily coordinator workload and projected time to reach a fixed enrollment target as functions of the decision threshold, assuming a 30 patient ICU census, 7 minutes per chart review, 60 minutes per consent attempt, and an 8 hour daily time budget (dashed line). Higher thresholds reduce review time but sharply increase days to enrollment due to missed eligible patients; the selected operating point (*t* ≈ 0.23, vertical line) balances feasibility and recruitment yield. (B) Matrix for the random forest model at *t* = 0.23, showing per-100-day true positives, false positives, true negatives, and false negatives, illustrating high capture of ICU-to-floor transfers at manageable review burden.

At the optimal operating point (threshold ≈ 0.23), the random forest model identified approximately 75.7 true ICU-to-floor transfers per 100 ICU patient days (Figure 6B), corresponding to ∼22.7 true transfers identified per day assuming an ICU census of 30 patients. Under conservative recruitment assumptions, 10% eligibility among true transfers and a 60% consent rate, this translated to approximately 1.36 enrolled patients per day. Total coordinator workload at this threshold was approximately 339 minutes per day, including ∼210 minutes for chart review and ∼136 minutes for approach and consent, remaining within the daily time budget. Under these assumptions, this operating point yielded an estimated net value of approximately $2,380 per day, reflecting enrollment driven gains that exceeded coordinator labor costs.

These findings are particularly relevant in the context of modern pharmaceutical trials, where personnel costs and trial duration are major drivers of overall expense Martin et al. (2017); Chaudhari et al. (2020); Gul and Ali (2010); Khanna (2012). Prior work has shown that even modest reductions in recruitment inefficiency or cycle time can translate into substantial cost savings at scale. Our results demonstrate that such gains are not achieved by maximizing predictive accuracy alone, but by selecting operating thresholds that align with the structure of recruitment funnels and staffing constraints. In this setting, avoiding missed eligible patients was more consequential than minimizing review effort, a trade-off that would be obscured without explicit decision analytic evaluation.

The interaction between recruitment efficiency and threshold selection is further illustrated by the projected time required to reach a fixed enrollment target of 300 patients, shown alongside workload constraints in Figure 6A. Across most thresholds, projected enrollment time ranged between approximately 200 and 400 days, indicating that recruitment speed is limited primarily by the rarity of eligible patients rather than screening capacity. Although higher thresholds reduce review burden, they substantially increase missed eligible ICU-to-floor transfers, which dominates net value under realistic recruitment funnels. When accounting jointly for screening time, consent time, eligibility rates, enrollment targets, and coordinator capacity, the optimal operating point shifts to a lower threshold (*t* ≈ 0.23), which captures nearly all eligible transfers while remaining operationally feasible.Overall, these results demonstrate that although multiple thresholds yield favorable net benefit under decision curve analysis, only a subset correspond to feasible and efficient clinical trial recruitment workflows. Incorporating explicit time, staffing, and enrollment constraints resolves ambiguity in threshold selection and converts discharge prediction outputs into a defensible recruitment prioritization policy. More broadly, these findings reinforce that the practical value of ICU discharge prediction models depends not on discrimination alone, but on how predictions are translated into actions under real operational constraints.

## 4. Discussion

Predictive models for ICU discharge have traditionally been evaluated as forecasting tools, with emphasis placed on discrimination and calibration metrics such as ROC AUC, PR AUC, and Brier score. While these metrics quantify statistical accuracy, they do not determine whether acting on model predictions improves real-world workflows when actions result in asymmetric and resource constrained costs. In this study, we reframed next day ICU discharge prediction as a decision support problem for clinical trial recruitment, where predictions are used to prioritize proactive chart review and patient outreach under fixed coordinator capacity. By applying decision curve analysis and extending it with an explicit recruitment funnel and time budget, we showed that the operational value of discharge predictions is driven less by ranking performance and more by how thresholds interact with eligibility constraints. In particular, when realistic eligibility (10%) and consent (60%) rates were accounted for, the optimal operating point shifted to a lower threshold (*t* ≈ 0.23), capturing nearly all eligible ICU-to-floor transfers while remaining feasible within an 8-hour workday and yielding an estimated net value of approximately $2,380 per day.

Across models, discrimination and precision– recall performance were similar (ROC AUC 0.80– 0.84; PR AUC *>* 0.91), yet decision curve analysis revealed threshold-dependent differences in operational utility that were not evident from these metrics alone. At low thresholds, model guided strategies achieved net benefit comparable to or exceeding review-all largely by flagging nearly all patient days, providing minimal reduction in review burden in this high prevalence setting. By explicitly linking probability thresholds to false positive penalties and downstream workload, DCA clarified when apparent gains reflected meaningful prioritization versus trivial sensitivity increases. These results show that, for ICU discharge prediction, favorable discrimination does not imply operational benefit unless threshold selection meaningfully alters review effort and missed eligible patients.

A key contribution of this work is the integration of DCA with a realistic clinical recruitment workflow. Although higher thresholds reduced chart review burden, they substantially increased missed eligible patients, which dominated net value under realistic recruitment funnels. When screening time was reduced to reflect practical review durations, the limiting factor was no longer workload but the rarity of eligible patients within the ICU population. As a result, lower thresholds that maximize sensitivity, while still remaining operationally feasible, were preferred. This finding highlights why threshold selection cannot be justified by predictive accuracy or DCA alone, but must be grounded in the structure of the downstream workflow the model is intended to support.

Framing prediction outputs as inputs to prioritization, rather than as automated clinical decisions, further distinguishes this work from prior ICU discharge studies. In our analysis, a positive prediction triggers anticipatory review or outreach, leaving final judgment to clinicians or research staff. This human-in-the-loop framing aligns with best practices for clinical decision support, where models are intended to augment rather than replace expert decision making Ad-lung et al. (2021); Schwartz et al. (2021). By contrast, much of the ICU discharge literature implicitly evaluates models as if improved accuracy directly translates into safer or earlier discharge decisions. Our results demonstrate that even well calibrated models may offer limited operational value if predictions are not evaluated in relation to the specific actions they inform.

Several prior studies have shown that ML models can predict ICU discharge readiness, length of stay, or readmission with reasonable accuracy. However, evaluation has largely centered on discrimination metrics or retrospective comparisons to clinician behavior, without explicitly assessing whether model use would improve workflows or resource allocation. Recent work in hospital operations has begun to emphasize the importance of linking predictions to operational out-comes, but decision analytic evaluation remains less common in ICU focused studies Pianykh et al. (2020); Bertsimas et al. (2022); Levin et al. (2021). Our findings extend this literature by demonstrating how DCA can be used not only to compare models, but to design and justify implementable recruitment policies. By making threshold dependent trade-offs explicit and tying them to staffing constraints and enrollment yield, this framework allows stakeholders to select operating points consistent with institutional priorities rather than abstract performance benchmarks.

This study has several limitations. All analyses were retrospective and conducted using data from a single health system, which may limit generalizability Kim et al. (2016); Angelo et al. (2017). Economic and recruitment parameters were treated as illustrative, though chosen to reflect realistic ICU trial conditions, and acceptable trade-offs between workload and missed opportunities would require stakeholder input in practice. Future work should focus on prospective, human-in-the-loop evaluation of model guided recruitment prioritization, comparing algorithmic and clinician driven workflows in real time. Such studies could quantify downstream effects on enrollment speed, coordinator burden, and trial completion, and assess generalizability across institutions.

Extending this decision analytic framework to other ICU prediction tasks may help standardize evaluation of machine learning models in high stakes clinical settings. In this study, we demonstrated that the practical value of ICU discharge prediction models depends not on discrimination alone, but on how predicted risks are translated into actions under realistic operational constraints. By combining decision curve analysis with explicit modeling of clinical trial recruitment workflows, we identified an implementable operating point that balanced sensitivity, workload, and enrollment yield in an eligibility limited setting. These findings highlight the importance of decision analytic evaluation for determining when and how machine learning models should be deployed to support real world clinical and research workflows.

## Data Availability

This research has been conducted using data from MIMIC-IV. Researchers can request access via PhysioNet.

https://physionet.org/content/mimiciv/3.1/

## 5. Citations and Bibliography

## Acknowledgments

This study was supported the Huntsman Mental Health Foundation, the National Institute of Aging of the National Institutes of Health under Award Number L70AG096751, and the University of Utah’s Digital Health Initiative (W.W.P.). National Heart, Lung, and Blood Institute of the National Institutes of Health under Award Number R01HL169521 and the Intermountain Fund under Project Number 20400306 (BWL).

## Appendix A First Appendix

**Table 2:** Threshold interpretation for decision curve analysis. For each threshold probability *t*, the implied relative penalty of a false positive decision (unnecessary review) versus a false negative decision (missed next-day ICU transfer) is 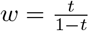. The interpretations translate this trade-off into an approximate number of unnecessary reviews considered acceptable to avoid missing one true next-day ICU transfer.

| Threshold ($t$) | FP penalty weight ($w$) | Interpretation |
| --- | --- | --- |
| 0.1 | 0.111 | Avoiding a missed ICU-to-hospital-floor transfer is considered more harmful than an additional review. |
| 0.3 | 0.429 | Performing approximately 0.4 unnecessary reviews is preferable to missing one patient transferring from ICU to hospital floor. |
| 0.5 | 1.000 | One unnecessary review is equally costly as missing one true patient transferring from ICU to hospital floor. |
| 0.7 | 2.333 | Missing one patient transferring from ICU to hospital floor is acceptable if it avoids more than two unnecessary reviews. |
| 0.9 | 9.000 | Missing one true ICU-to-hospital-floor transfer is acceptable if it avoids nine unnecessary reviews. |

## References

Lorenz Adlung, Yotam Cohen, Uria Mor, and Eran Elinav. Machine learning in clinical decision making, 6 2021. ISSN 26666340.

Simone A. Angelo, Edilson F. Arruda, Rosane Goldwasser, Maria S.C. Lobo, André Salles, and José Roberto Lapa e Silva. Demand forecast and optimal planning of intensive care unit (icu) capacity. Pesquisa Operacional, 37:229–245, 5 2017. ISSN 16785142. doi: 10.1590/0101-7438.2017.037.02.0229.

Tony Antoniou and Muhammad Mamdani. Evaluation of machine learning solutions in medicine. CMAJ, 193:E1425–E1429, 9 2021. ISSN 14882329. doi: 10.1503/cmaj.210036.

Omar Badawi and Michael J. Breslow. Readmissions and death after icu discharge: Development and validation of two predictive models. PLoS ONE, 7, 11 2012. ISSN 19326203. doi: 10.1371/journal.pone.0048758.

Dimitris Bertsimas, Jean Pauphilet, Jennifer Stevens, and Manu Tandon. Predicting inpatient flow at a major hospital using interpretable analytics. Manufacturing and Service Operations Management, 24:2809–2824, 11 2022. ISSN 15265498. doi: 10.1287/msom.2021.0971.

Nayan Chaudhari, Renju Ravi, Nithya Gogtay, and Urmila Thatte. Recruitment and retention of the participants in clinical trials: Challenges and solutions. Perspectives in Clinical Research, 11:64–69, 4 2020. ISSN 22295488. doi: 10.4103/picr.PICR_206_19.

Cory Franklin and David Jackson. Jackson decision making icu. Critical Care Medicine, 11, 2 1983.

Raisa B. Gul and Parveen A. Ali. Clinical trials: The challenge of recruitment and retention of participants. Journal of Clinical Nursing, 19: 227–233, 1 2010. ISSN 09621067. doi: 10.1111/j.1365-2702.2009.03041.x.

Teus H. Kappen, Wilton A. van Klei, Leo van Wolfswinkel, Cor J. Kalkman, Yvonne Vergouwe, and Karel G. M. Moons. Evaluating the impact of prediction models: lessons learned, challenges, and recommendations. Diagnostic and Prognostic Research, 2, 12 2018. doi: 10.1186/s41512-018-0033-6.

Ish Khanna. Drug discovery in pharmaceutical industry: Productivity challenges and trends, 10 2012. ISSN 13596446.

Song Hee Kim, Carri W. Chan, Marcelo Olivares, and Gabriel J. Escobar. Association among icu congestion, icu admission decision, and patient outcomes. Critical Care Medicine, 44:1814–1821, 10 2016. ISSN 15300293. doi: 10.1097/CCM.0000000000001850.

Scott Levin, Sean Barnes, Matthew Toerper, Arnaud Debraine, Anthony Deangelo, Eric Hamrock, Jeremiah Hinson, Erik Hoyer, Trushar Dungarani, and Eric Howell. Machine-learning-based hospital discharge predictions can support multidisciplinary rounds and decrease hospital length-of-stay. BMJ Innovations, 7:414–421, 4 2021. ISSN 2055642X. doi: 10.1136/bmjinnov-2020-000420.

Linda Martin, Melissa Hutchens, Conrad Hawkins, and Alaina Radnov. How much do clinical trials cost?, 6 2017. ISSN 14741784.

Oleg S. Pianykh, Steven Guitron, Darren Parke, Chengzhao Zhang, Pari Pandharipande, James Brink, and Daniel Rosenthal. Improving healthcare operations management with machine learning. Nature Machine Intelligence, 2:266–273, 5 2020. ISSN 25225839. doi: 10.1038/s42256-020-0176-3.

Juan C. Rojas, Kyle A. Carey, Dana P. Edelson, Laura R. Venable, Michael D. Howell, and Matthew M. Churpek. Predicting intensive care unit readmission with machine learning using electronic health record data. Annals of the American Thoracic Society, 15:846–853, 7 2018. ISSN 23256621. doi: 10.1513/AnnalsATS.201710-787OC.

Hanna Bloomfield Rubins and Mark Moskowitz. Discharge decision-making in a medical intensive care unit identifying patients at high risk of unexpected death or unit readmission. Technical report, Boston University Medical Center, 5 1988.

Jessica M. Schwartz, Amanda J. Moy, Sarah C. Rossetti, Noémie Elhadad, and Kenrick D. Cato. Clinician involvement in research on machine learning-based predictive clinical decision support for the hospital setting: A scoping review, 3 2021. ISSN 1527974X.

Ashley Siems, Russell Banks, Richard Holubkov, Kathleen L. Meert, Christian Bauerfeld, David Beyda, Robert A. Berg, Yonca Bulut, Randall S. Burd, Joseph Carcillo, J. Michael Dean, Eleanor Gradidge, Mark W. Hall, Patrick S. McQuillen, Peter M. Mourani, Christopher J.L. Newth, Daniel A. Notterman, Margaret A. Priestley, Anil Sapru, David L. Wessel, Andrew R. Yates, Athena F. Zuppa, and Murray M. Pollack. Structured chart review: Assessment of a structured chart review methodology. Hospital Pediatrics, 10:61–69, 1 2020. ISSN 21541671. doi: 10.1542/hpeds.2019-0225.

Michelle Tew, Max Catchpool, John Furler, Katie De La Rue, Philip Clarke, Jo Anne Manski-Nankervis, and Kim Dalziel. Site-specific factors associated with clinical trial recruitment efficiency in general practice settings: a comparative descriptive analysis. Trials, 24, 12 2023. ISSN 17456215. doi: 10.1186/s13063-023-07177-4.

Ilona Willempje Maria Verburg, Alireza Atashi, Saeid Eslami, Rebecca Holman, Ameen Abu-Hanna, Everet De Jonge, Niels Peek, and Nicolette Fransisca De Keizer. Which models can i use to predict adult icu length of stay? a systematic review, 2 2017. ISSN 15300293.

Andrew J. Vickers and Ford Holland. Decision curve analysis to evaluate the clinical benefit of prediction models. Spine Journal, 21:1643–1648, 10 2021. ISSN 18781632. doi: 10.1016/j.spinee.2021.02.024.

Andrew J. Vickers, Ben van Calster, and Ewout W. Steyerberg. A simple, step-by-step guide to interpreting decision curve analysis. Diagnostic and Prognostic Research, 3, 12 2019. ISSN 2397-7523. doi: 10.1186/s41512-019-0064-7.

